# Efficacy and Safety of Total Body Irradiation versus Chemotherapy Conditioning for Hematopoietic Stem Cell Transplant in Adult Acute Lymphoblastic Leukemia: A Systematic Review and Meta-Analysis

**DOI:** 10.1101/2024.11.16.24316211

**Authors:** Hasaan Ans, Haiqah Amjad, Sara Nazir, Sara Sajjad Cheema, Dhruv Mistry, Tazeen Fatima, Ahmed Yar Khan, Muhammad Usama Imtiaz, Fasih Mand Khan, Saif Khalid, Mohammad Ebad Ur Rehman, Muhammad Salman Faisal

## Abstract

**Background:** Hematopoietic stem cell transplantation (HSCT) is a potentially curative option for adults with acute lymphoblastic leukemia (ALL) who have achieved remission. This systematic review and meta-analysis compare the efficacy of total body irradiation (TBI) versus chemotherapy (CHT) based regimens for conditioning in adult ALL patients being prepared for HSCT.

**Methods:** A comprehensive literature search was conducted in MEDLINE, Embase, the Cochrane Library, and relevant trial registries from their inception to August 2024. The inclusion criteria encompassed all randomized controlled trials (RCTs) and cohort studies that compared TBI) with CHT as conditioning regimens prior to HSCT in adult patients with ALL. The primary outcomes assessed were overall survival (OS) and event-free survival (EFS). All statistical analyses were carried out using RevMan version 5.4, using a random-effects model.

**Results:** This meta-analysis includes 20 cohort studies and one RCT. The relative risk (RR) for OS was 1.37 (95% CI: 1.20–1.57), while the RR for EFS was 1.28 (95% CI: 1.15–1.43), highlighting the superior efficacy of TBI-based regimens in this patient population. TBI was also superior in terms of relapse rate (RR 0.71). The two regimens were comparable in terms of non-relapse mortality, acute graft-versus-host disease (GVHD), and chronic GVHD.

**Conclusion:** When used for conditioning prior to HSCT, TBI-based conditioning regimens demonstrate superior OS, EFS, and relapse outcomes compared to CHT-based regimens.

## Introduction

Acute lymphoblastic leukemia (ALL), a malignant disorder involving lymphoid progenitor cells, manifests in both pediatric and adult populations, with the highest incidence observed between 2 and 5 years of age^1^. Significant progress in treatment development has achieved cure rates above 80% in children. The use of risk-based stratification and enhanced chemotherapy protocols has greatly improved survival rates in those with ALL, especially in pediatric patients (ages 1-14) and also in adolescents and young adults (ages 15-39). Despite these advances, outcomes for older adults (≥40 years) and those with relapsed or refractory ALL remain unsatisfactory^1,2^. Standard management of ALL typically involves a combination of chemotherapy administered in phases of induction, consolidation, and maintenance spanning 2.5 to 3 years. This regimen also includes prophylactic measures for the central nervous system (CNS) and may incorporate allogeneic stem cell transplantation (ASCT) during initial or subsequent remissions^3^. Transplantation, compared to standard methods, may lead to improved disease-free survival rates. However, the success of this approach is significantly influenced by patient selection criteria. It is recommended for pediatric patients with ALL who have relapsed or are at high risk of relapse during their first complete remission, as well as for patients who fail to achieve MDR negativity, have not received intensive induction therapy, or exhibit high-risk cytogenetic and molecular characteristics^4–6^.

In patients with ALL, prior to allogeneic and autologous hematopoietic stem cell transplantation (HSCT), total body irradiation (TBI) is administered as part of the conditioning regimen, delivering a relatively homogeneous dose of radiation to the entire body. TBI functions in two capacities: it is both cytotoxic and immunosuppressive. This eliminates the remaining disease and creates space in the bone marrow, weakening the immune system to prevent rejection of transplanted donor cells^7^. There is ongoing debate over whether chemotherapy (CHT) or TBI is more effective as part of conditioning regimens for patients undergoing HSCT. TBI has certain advantages over chemotherapy, such as greater tumor cytotoxicity and the ability to penetrate tissues more effectively. Since its administration does not rely on blood flow or physiological barriers like renal or hepatic function, TBI can reach “sanctuary” regions, including the CNS, testes, and orbits, which may be less accessible to CHT^7^.

Several meta-analyses have evaluated TBI versus CHT-based conditioning before HSCT in adults with ALL, with promising results showing a statistically significant reduction in side effects and an improvement in overall survival (OS). However, new studies have since been published. A pooled analysis could provide further insight into critical outcomes such as graft-versus-host disease (GVHD), relapse, mortality, and event-free survival (EFS). Therefore, we have conducted a systematic review and meta-analysis to compare outcomes between TBI and CHT conditioning in the adult ALL population.

## Methods

Our meta-analysis was performed following the guidelines outlined in the Cochrane Handbook for Systematic Reviews of Interventions, and the results are reported in line with the Preferred Reporting Items for Systematic Reviews and Meta-Analyses (PRISMA) statement^8,9^. Our protocol is registered with PROSPERO, the International Prospective Register of Systematic Reviews.

### Data sources and Searches

We conducted an electronic search of the Cochrane Central Register of Controlled Trials (CENTRAL, via the Cochrane Library), MEDLINE, Embase (Elsevier), and ClinicalTrials.gov from their inception through August 2024 using a comprehensive search strategy. MeSH terms and keywords for “Whole Body Irradiation” and “Precursor Cell Lymphoblastic Leukemia-Lymphoma” were included. Additionally, we reviewed reference lists of included studies and related systematic reviews to identify other relevant studies. Detailed search strategy for Pubmed (MEDLINE) is reported in Supplementary Table 1.

### Eligibility Criteria

The inclusion criteria were: (1) population: adult patients with ALL, (2) intervention: TBI-based regimens, (3) comparison: CHT-based regimens, and (4) study design: RCTs or cohort studies. The exclusion criteria were: (1) any study designs other than RCTs and cohort studies and (2) studies involving animals or minors.

### Study Selection and Data Abstraction

We imported all studies identified through our online search into Rayan and removed duplicates. Two authors (H.A. and D.M.) independently screened the titles and abstracts, excluding irrelevant articles. We then conducted a full-text review of the remaining studies and finalized those that met our eligibility criteria. A third author (M.E.U.R.) resolved any disagreements during the study selection process.

Two review authors (H.A. and M.U.I.) extracted data from the included studies using a pre-piloted data extraction form into an Excel spreadsheet. Data pertaining to author name, year of publication, study setting, study design, sample size, age, ALL subtype, remission status, stem cell source, dose, frequency an duration of TBI and CHT regimens used were extracted. The extracted data were compared, and any discrepancies were addressed through discussion with a third author (M.E.U.R.).

### Outcome Measures

The primary outcome measures were OS and EFS. The secondary outcome measures were relapse, non-relapse mortality (NRM), acute graft versus host disease (aGVHD), acute graft versus host disease grade 3 and 4 (aGVHD 3-4) and chronic graft versus host disease (cGVHD).

### Risk of Bias Assessment

Two independent investigators (S.N. and A.Y.A.) assessed the quality of the included RCT and cohort studies using the Revised Cochrane Risk of Bias tool (RoB 2.0) and the Newcastle-Ottawa Scale (NOS), respectively. Any discrepancies were resolved by a senior investigator (M.E.U.R.).^10,11^.

### Data Synthesis

Review Manager (RevMan, Version 5.4) performed the meta-analyses. Risk ratios (RRs) and corresponding 95% confidence intervals (CIs) for both primary and secondary outcomes were extracted from each study. A random-effects model with the Mantel-Hanszel method was employed for the meta-analyses. Statistical heterogeneity was assessed using the Higgins I² statistic.

## Results

### Search Results

The initial search garnered 1,012 articles. Title and abstract screening resulted in the exclusion of 854 articles, and 61 out of the remaining 82 studies were excluded after full-text screening. Finally, 21 articles were included in this meta-analysis. The detailed screening process is depicted in Figure 1.

**Figure 1:**
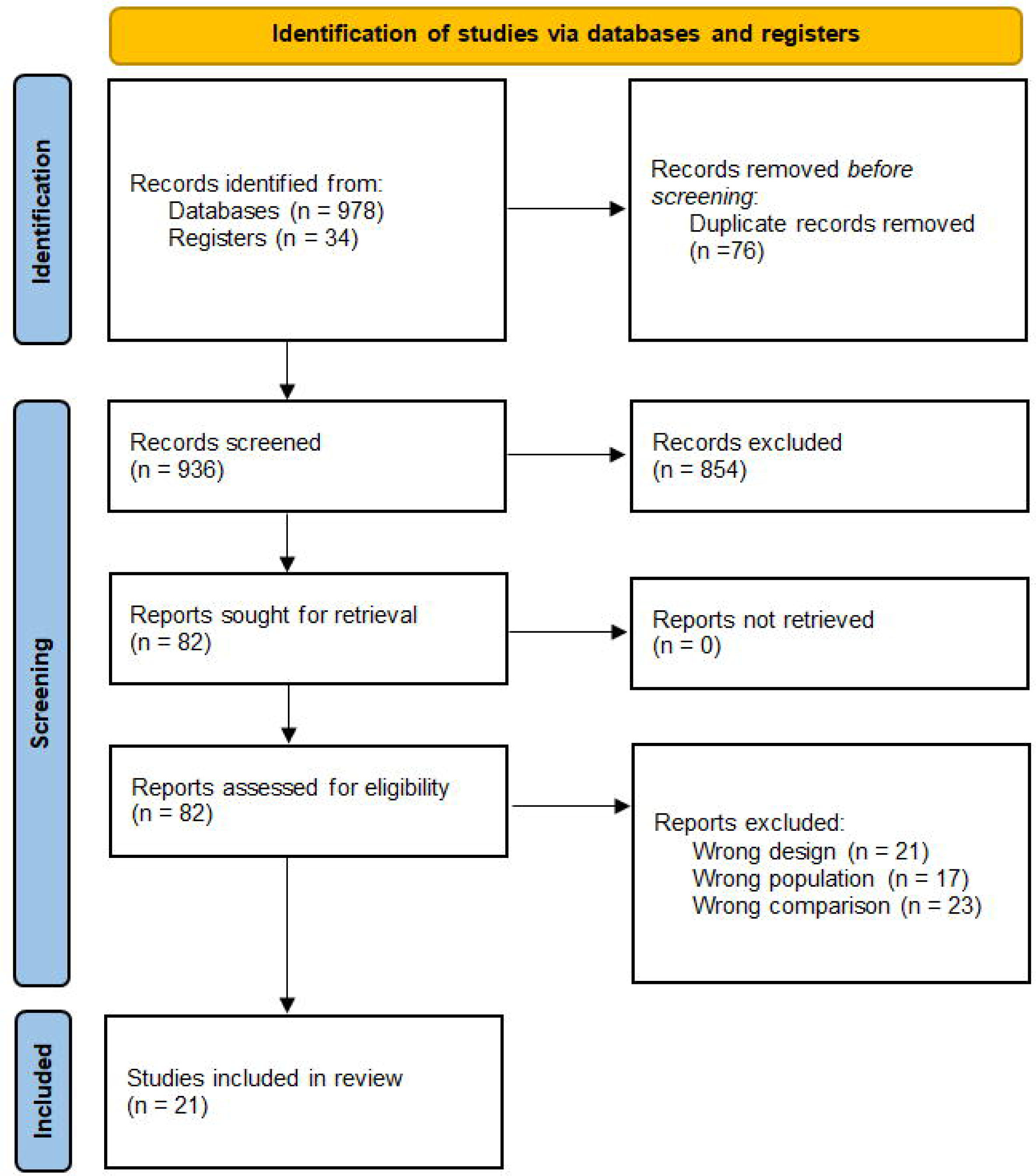
PRISMA Flowchart.

### Study Characteristics

A total of 21 studies were included in this meta-analysis^12–32^. The studies comprised eleven cohort studies and one RCT. Sample sizes ranged from 95 to 2,780 patients, with a cumulative total of 12,046 participants. Conditioning regimens were TBI-based, typically combined with etoposide or cyclophosphamide, or CHT-based regimens, which commonly included busulfan, fludarabine, and cytarabine. Detailed study characteristics, including sample sizes, patient demographics, and conditioning protocols, are provided in Table 1.

**Table 1:** Characteristics of Included Studies.

| Study ID | Setting | Study Design | Sample Size | Mean Age, in years (S.D.) | ALL Subtype (B-cell/T-cell) | Remission Status (CR 1/Advanced) | Stem Cell Source (BM/PS C/CB) | TBI Regimen | CHT Regimen |
| --- | --- | --- | --- | --- | --- | --- | --- | --- | --- |
| Abdeljelil 2024 | Tunisia | Retrospective Cohort | 95 ( 61 vs 34 ) | 30 (18-50)* | 56/39 | 80/15 | 46/49/0 | TBI 9.9 Gy + Etoposide (60 mg/kg per day)/Cyclophosphamide (60 mg/kg per day) | Busulfan 3.2 mg/kg per day and Cyclophosphamide 60mg/kg for two days (Bu-Cy) or Fludarabine 30 mg/m2 once a day for 6 days and Busulfan 3.2 mg/kg per day and Cyclophosphamide 60mg/kg for two days (FBC) or Thiotepa 5 mg/kg once a day for 2 days and Busulfan 3.2 mg/kg per day and Fludarabine 50 mg/m2 once a day for 3 days (TBF). |
| Bazarbachi | Europe | Retrospective | 115 (60 vs 55) | 30 (18- | 35/34 | 0/115 | NR | 8 Gy | Oral Busulfan |
| 2020 |  | Cohort |  | 63)* |  |  |  |  | greater than 8 mg/kg or IV Bu greater than 6.4 mg/kg |
| Cahu 2015 | Europe | Retrospective Cohort | 601 ( 523 vs 78 ) | 30 ( 18-63 ) * | 0/601 | 414/187 | 196/405/0 | TBI and Cyclophosphamide | IV busulfan–cyclophosphamide |
| Dholaria 2021 | Europe | Retrospective Cohort | 427 ( 188 vs 239 ) | 31* | 291/136 | 208/219 | 229/198/0 | TBI and Fludarabine | thiotepa–busulfan–fludarabine |
| Eroglu 2013 | Turkey | Retrospective Cohort | 95 ( 45 vs 50 ) | 25 (9–54)* | NR | 81/14 | NR | TBI 12 Gy + Cyclophosphamide 60 mg/kg/day | Busulfan 3.2 mg/kg/day and Cyclophosphamide 60 mg/kg/day |
| Greil 2020 | Germany | Retrospective Cohort | 180 (119 vs 61) | 37 (16–76)* | 50/44 | 38/41 | 27/153/0 | TBI 12 Gy | NR |
| Harada 2019 | Japan | Retrospective Cohort | 507 (355 vs 152) | HDM/TBI 57 (16–69) / LDM/TBI 61 (21–71) vs HDM 58 (16–67)* | 417/27 | 388/72 | 237/72/198 | TBI 2–4 Gy and Melphalan (20–140 mg/m2 | Melphalan 120–140mg/m2 |
| Hirschbühl 2023 | Europe | Retrospective Cohort | 501 (262 vs 239) | FluTB I8 56 (45.4–76.1)/ FluBu 6.4 60.3 (45.1–72)/ | 435/66 | NR | 32/469/0 | TBI 8 Gy and Fludarabine | Fludarabine and busulfan 6.4/9.6 mg/kg |
|  |  |  |  | FluBu<br>9.6<br>55.4<br>(45.9–<br>65.6)* |  |  |  |  |  |
| Kalaycio<br>2010 | USA | Retrospective<br>Cohort | 86 (35 vs<br>51) | 40<br>(19-56) vs<br>33<br>(19-60)* | 65/14 | 35/47 | 78/4/0 | TBI 1200cGy<br>+ Etoposide 60<br>mg/kg | Busulfan 16<br>mg/kg,<br>Cyclophosphamide 120<br>mg/kg or<br>Busulfan 16<br>mg/kg,<br>Cyclophosphamide 120<br>mg/kg,<br>Etoposide<br>50mg/kg |
| Kebriaei<br>2018 | USA | Retrospective<br>Cohort | 1118<br>(819 vs<br>299) | 37<br>(18-60) vs<br>38<br>(18-60)* | 924/1<br>94 | 833/28<br>5 | 244/874/<br>0 | TBI 9-12 Gy +<br>Cyclophosphamide 106 (89.5<br>-119.5) mg/kg<br>or TBI 13-16Gy +<br>Etoposide 54<br>(47-59) mg/kg | Busulfan<br>11.5 (9.8-12.8) mg/kg<br>and<br>Clolarabine<br>160mg/m2,<br>or Busulfan<br>11.5 (9.8-12.8) mg/kg<br>and<br>Fludarabine<br>160 (159-160)<br>mg/m2, or<br>Busulfan<br>11.5 (9.8-12.8) mg/kg<br>and<br>Melphalan<br>(129-140)<br>140mg/m2,<br>or Busulfan<br>11.5 (9.8-12.8) mg/kg<br>and<br>Cyclophosphamide<br>120mg/kg |
| Konuma<br>2022 | Japan | Retrospective Cohort | 333 (258 vs 75) | 41 (16–61) 52 (19–69)* | NR | NR | 0/0/333 | TBI 8-12 Gy, Cyclophosphamide 120 mg/kg, highdose Cytosine Arabinoside (Ara-C) 12 g/m2 | Busulfan 9.6 mg/kg, Fludarabine 180 mg/m2, high-dose Ara-C 12 g/m2, or Busulfan 12.8 mg/kg, Cyclophosphamide 120 mg/kg, Fludarabine 100 mg/m2 |
| Mora<br>2024 | Spain | Retrospective Cohort | 177 (114 vs 63) | 31 (18–53) vs 38 (18–56) | 138/39 | 101/76 | 13/164 | TBI 12-13Gy + Cyclophosphamide 120mg/kg or TBI 12-13Gy + Fludarabine 120mg/m2 | Thiotepa 10mg/kg, Busulfan 9.6mg/kg, Cyclophosphamide 120mg/kg or Thiotepa 10mg/kg, Busulfan 9.6mg/kg, Fludarabine 120mg/m2 |
| Mutsuhashi<br>2016 | Japan | Retrospective Cohort | 2130 (2028 vs 102) | 39 (16-68)* | NR | 1771/359 | 1400/333/397 | TBI ≥8Gy + Cyclophosphamide 120mg/kg | Busulfan 16 mg/kg, Cyclophosphamide 120 mg/kg, or Busulfan 12.8 mg/kg, Cyclophosphamide 120 mg/kg |
| Niu<br>2022 | China | Retrospective Cohort | 40 (23 vs 17) | 28 (18-46) vs 28.5 (18-50)* | 17/23 | 34/6 | 0/40/0 | TBI 10Gy + Cyclophosphamide 50-60mg/kg+ Etoposide 15 mg/kg | Busulfan 3.2 mg/Kg, Cyclophosphamide 50-60mg/Kg |
| Park<br>2019 | Korea | Retrospective | 1562(735 vs | 39 (18- | NR | 482/1080 | NR | Cyclophosphamide (524/735, | Busulfan or Fludarabine |
|  |  | Cohort | 827) | 65) vs<br>41<br>(18-65)* |  |  |  | 71.2%), high-dose cytarabine<br>(199/735,<br>27.1%), or<br>melphalan<br>(12/735, 1.6%) | and<br>Busulfan |
| Pavlu<br>2019 | Euro<br>pe | Registry<br>Study | 2780 (<br>2122 vs<br>658) | MRD<br>+38<br>(18-72,<br>28-48),<br>MRD-36<br>(18-70,<br>26-46) | 2181/<br>599 | 2427/3<br>53 | 670/211<br>0/0 | NR | NR |
| Sakella<br>ri 2018 | Gree<br>ce | Retrospe<br>ctive<br>Cohort | 151 (84<br>vs 67) | 29.4(1<br>7-41.6)<br>vs<br>29.1<br>(16.4-41.8) | 114/3<br>7 | 60/89 | 26/125/0 | TBI 14.4Gy | Busulfan 4<br>mg/Kg |
| Shen<br>2024 | Chin<br>a | Retrospe<br>ctive<br>Cohort | 222(<br>83vs<br>139) | 21 (4-49)<br>vs. 24<br>(2-52) | NR | 117/<br>105 | NR | TBI plus<br>Cyclophospha<br>mide | busulfan<br>plus<br>cyclophosp<br>hamide |
| Swobo<br>da<br>2022 | Euro<br>pe | Retrospe<br>ctive<br>Cohort | 237(117v<br>s119) | 30.9<br>(18.1-59.9)<br>vs<br>35.8<br>(18.8-61.7) | 171/6<br>5 | 128/10<br>9 | 141/95 | TBI >10 Gy<br>plus<br>Fludarabine | Thiotepa<br>(two days,<br>total dose<br>of 10<br>mg/kg),<br>Busulfan<br>(three days,<br>total dose<br>of 9.6<br>mg/kg) and<br>Fludarabine |
| Wang<br>2021 | Taiw<br>an | Retrospe<br>ctive<br>Cohort | 224 (83<br>vs 95) | 28.1<br>(16.4-<br>56.3)<br>vs<br>30.9<br>(15.5- | 137/6<br>5 | 114/11<br>0 | 50/159/ | TBI 150 centi-Gy and<br>Cyclophospha<br>mide 60<br>mg/kg/day | busulfan IV<br>3.2<br>mg/kg/day<br>or oral 4<br>mg/kg/day<br>PLUS |
|  |  |  |  | 52.3)* |  |  |  |  | cyclophosphamide IV 60 mg/kg/<br>OR<br>fludarabine 30 mg/m2/day , busulfan IV 3.2 mg/kg/day or oral 4 mg/kg/day and cyclophosphamide IV 60 mg/kg/day |
| Zhang 2022 | China | Randomized Control Trial | 545 ( 272 vs 273 ) | 27 (14-61) vs 26 (14-59) | NR | NR | NR | 4.5 Gy/day plus cyclophosphamide 60 mg/kg once daily | Busulfan 3.2 mg/kg per day with cyclophosphamide 60 mg/kg per day |

### Risk of Bias in Included Studies

The quality assessment of the included studies is presented in Supplementary Figure 1 and Supplementary Table 2. The RCT by Zhang et al. demonstrated a low risk of bias across all domains. Out of the twenty observational studies, five had a score of 9, seven had a score of 8, six had a score of 7, and two had a score of 6.

## Synthesis of Primary Outcomes

### Overall Survival

The meta-analysis for OS included data from 18 studies. The pooled results favoured TBI over CHT, with a RR of 1.37 (95% CI: 1.20–1.57, p<0.00001, I^2^ = 83%) (Figure 2).

**Figure 2:**
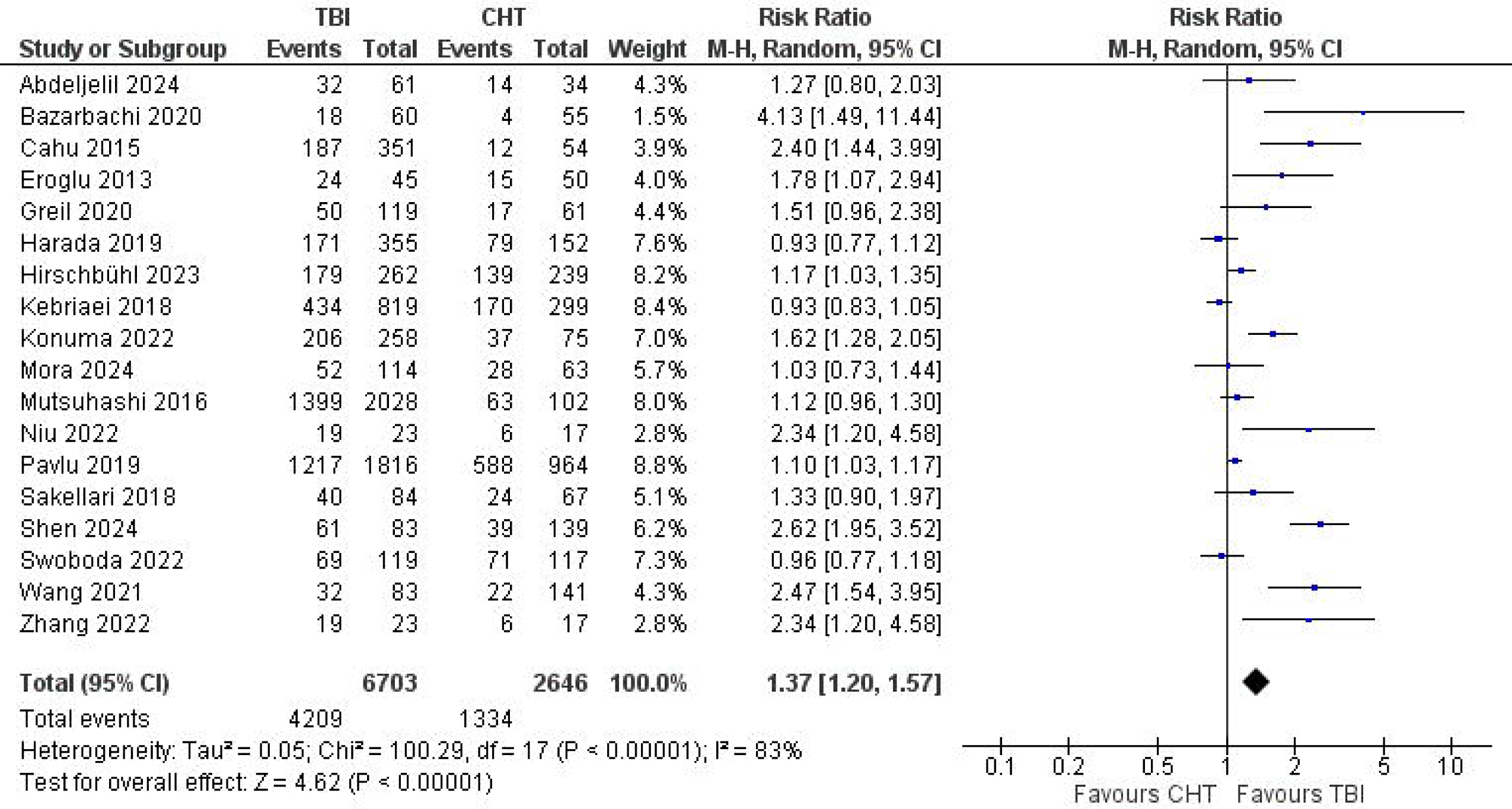
Forest Plot of Overall Survival.

### Event-Free Survival

EFS was analyzed across 17 studies. The pooled RR for EFS was 1.28 (95% CI: 1.15–1.43, p<0.00001, I^2^ = 45%), indicating a significant improvement in event-free survival for patients treated with TBI compared to CHT (Figure 3).

**Figure 3:**
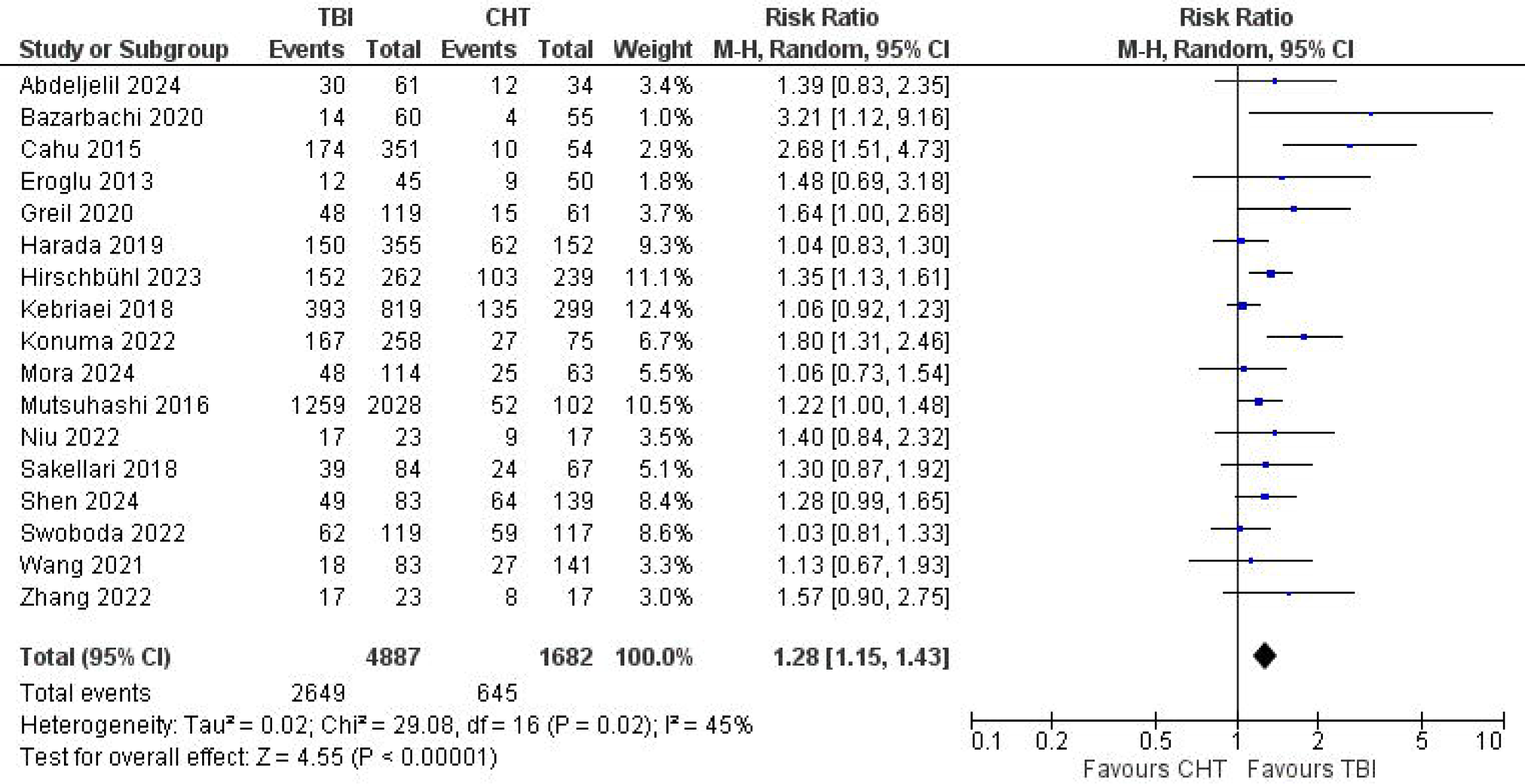
Forest Plot of Event-Free Survival.

### Relapse

Relapse was reported in 17 studies. The pooled RR for relapse was 0.71 (95% CI: 0.62–0.80, p<0.00001, I² = 63%), indicating a statistically significant reduction in relapse risk for patients receiving TBI compared to those receiving CHT (Supplementary Figure 2).

### Non-Relapse Mortality

NRM was evaluated in 17 studies. The pooled analysis showed no significant difference between TBI and CHT, with an RR of 0.86 (95% CI: 0.69–1.07, p = 0.17, I² = 69%) (Supplementary Figure 3).

### Acute Graft-Versus-Host Disease

The incidence of aGVHD was reported in 8 studies. The pooled RR was 1.02 (95% CI: 0.92– 1.13), with no statistically significant difference between TBI and CHT (p = 0.68). Heterogeneity was low (I² = 19%) (Supplementary Figure 4). aGVHD grades 3–4 was analyzed in 8 studies. The RR was 0.93 (95% CI: 0.66–1.31, p = 0.67, I² = 20%) with no significant difference between TBI and CHT (Supplementary Figure 5).

### Chronic Graft-Versus-Host Disease

cGVHD was reported in 10 studies. The pooled RR was 1.12 (95% CI: 0.95–1.31, p = 0.18, I² = 40%), indicating no significant difference between the two groups (Supplementary Figure 6).

## Discussion

Our meta-analysis, which included 20 cohort studies and 1 randomized controlled trial involving 12,046 patients, found that TBI as a conditioning regimen for ALL patients undergoing HSCT is associated with improved OS, EFS, and lower relapse rates than CHT-only myeloablative regimens. No significant differences were observed between the two regimens regarding NRM, aGVHD, aGVHD grades 3-4, or cGVHD.

Our findings show that TBI significantly enhances OS in ALL patients compared to CHT, with a relative risk (RR) of 1.37. EFS was also better with TBI, with an RR of 1.28. These results are consistent with prior research. For example, Khimani et al. analyzed eight observational, non-randomized retrospective studies involving 5,328 patients and reported that TBI-based regimens were associated with improved EFS compared to CHT-only regimens, with hazard ratios (HR) of 0.76 (p=0.002) and 0.74 (p=0.003)^33^. Similarly, Rehman et al. reviewed data from 15 studies with 5,629 pediatric patients and found that TBI-based regimens led to better OS and EFS, with relative risks (RR) of 1.21 and 1.34, respectively^6^. Eroglu et al. compared 95 patients undergoing HSCT with TBI plus cyclophosphamide (TBI + Cy) versus busulfan plus cyclophosphamide (Bu + Cy) and found that the median OS was 37 months for the TBI + Cy group compared to 12 months for the Bu + Cy group (p=0.003), with median EFS of 13 months versus 4 months (p=0.006), indicating a significant advantage for the TBI + Cy regimen^34^. Collectively, these studies support our conclusion that TBI-based conditioning is more effective than chemotherapy in adult ALL patients undergoing HSCT. In contrast, a retrospective study by Abdeljelil et al., involving 95 patients with 61 receiving TBI-based regimens and 34 receiving chemotherapies, found that TBI-based regimens were associated with better OS and EFS (52% vs. 42.6% and 49% vs. 34.6%, respectively). However, these results were not statistically significant, with p-values of 0.2 for both OS and EFS, likely due to the small sample size^12^.

Cytogenetic abnormalities are independent prognostic factors in ALL. The most important chromosomal abnormality in ALL is the Philadelphia chromosome (Ph), characterized by the balanced translocation t(9;22) (q34;q11). Other major cytogenetic abnormalities include t(4;11) (q21;q23) involving the MLL gene, translocations such as t(8;14), t(1;19), and t(10;14), and structural abnormalities such as 9p, 6q, and 12p^35^. Ph can also be detected by the polymerase chain reaction for the BCR-ABL fusion protein and is present in 20% to 30% of adults with ALL^35^. Ph positivity confers a uniformly poor prognosis with standard chemotherapy. The initial white blood cell (WBC) count at diagnosis is an important prognostic factor reported in every study of ALL^35^. An arbitrary cutoff of 30 × 10^9 WBCs/L for B-lineage ALL or 100 × 10^9 WBCs/L for T-lineage ALL has often been used in clinical studies. Understanding these mechanisms underscores the need for effective conditioning regimens and targeted therapies in ALL treatments.

Our meta-analysis also found that the incidence of adverse events, including cGVHD, was comparable between TBI and chemotherapy regimens. Khimani et al. reported no significant difference in cGVHD incidence between TBI-based and CHT-only regimens, with a risk ratio of 1.10 and a p-value of 0.63^33^. Similarly, Gooptu et al. found that the cumulative incidence of cGVHD was comparable between regimens, with a 2-year estimate of 55% for FluBu and 49% for CyTBI, and a p-value of 0.21^36^. Our study’s finding of a decreased relapse rate with TBI, showing an RR of 0.71 and a p-value of 0.0003, aligns with previous research. Khimani et al. similarly reported a lower incidence of relapse with TBI-based regimens compared to chemotherapy-only regimens, with an RR of 0.82 and a p-value of 0.02^33^. Additionally, Elvira Mora et al’s study, spanning from 2002 to 2018, compared 63 patients receiving TTB-based regimens with 114 patients receiving TBI-based regimens, finding a lower 5-year cumulative incidence of relapse in the TBI cohort (30%) compared to the TTB cohort (47%), with a p-value of 0.03^37^.

TBI offers several advantages when comparing TBI versus chemotherapy (CHT) as conditioning regimens for ALL patients before HSCT. The benefit of TBI is that it provides uniformity in the radiation dosage to the entire body and penetrates areas such as the central nervous system (CNS) and testes, which are barriers to chemotherapy. Chemotherapy, which uses agents like cyclophosphamide and busulfan to create DNA cross-links and induce cell death, may not provide the same comprehensive disease control as TBI. The effectiveness of chemotherapy can be inconsistent depending on the specific drugs and patient characteristics, potentially resulting in higher relapse rates. TBI delivers a high dose of ionizing radiation, causing widespread DNA damage and cell death in the hematopoietic system by inducing both single- and double-strand breaks in DNA. TBI is both myeloablative and immunosuppressive at high doses, but at lower doses, it is only immunosuppressive and not myeloablative. Non-myeloablative and reduced-intensity conditioning regimens use lower doses of TBI, focusing on immunosuppression only.

When TBI is combined with myeloablative conditioning regimens, it can be delivered before or after the chemotherapy, most often cyclophosphamide. The most common TBI schedules include twice-daily 2-Gy fractions given over 3 days (total dose 12 Gy); twice-daily 1.5-Gy fractions over 4-4.5 days (total dose 12-13.5 Gy); three-times-daily 1.2-Gy fractions over 4 days (total dose 12-13.2 Gy); and once-daily 3-Gy fractions for 4 days (total dose 12 Gy)^38^. The most common early side effects of TBI are fatigue and skin changes^39^. TBI has limited long-term complications in adults. Functional lung abnormalities are observed in 19% of cases, always without clinical effect. Clinical hypothyroidism is a very rare complication, and thyroid hormones are perturbed in 7%^40^. While chemotherapy might be less effective in disease control, it generally has fewer long-term complications than TBI.

This meta-analysis has several limitations. The retrospective studies introduce potential selection bias and variability in how patients were assigned to conditioning regimens. Differences in clinical practices, including TBI dosing and protocols, complicate the interpretation of results. The study also faces challenges due to inconsistent outcome definitions and incomplete long-term data on quality of life and post-transplant complications. Unmeasured confounders, such as genetic factors and previous treatments, may also influence the findings. The short follow-up period limits the assessment of long-term adverse effects. To address these issues, future research should focus on large-scale, prospective studies with standardized methodologies and extended follow-up to accurately evaluate and compare the efficacy and safety of TBI versus chemotherapy regimens.

In conclusion, our meta-analysis indicates that TBI as a conditioning regimen for ALL patients undergoing HSCT results in better OS, EFS, and reduced relapse rates compared to CHT-only regimens. No significant differences were observed between TBI and chemotherapy regarding NRM, and acute or chronic GVHD. However, due to limitations such as variability in treatment protocols and a short follow-up period, further large-scale research is needed to confirm these findings and optimize conditioning strategies for ALL patients.

## Data Availability

All data produced in the present study are available upon reasonable request to the authors

## Acknowledgments

Not applicable.

## Availability of data

The data supporting this study’s findings are available from the corresponding author upon reasonable request.

