## Supplementary material for "Efficacy and Safety of Total Body Irradiation versus Chemotherapy Conditioning for Hematopoietic Stem Cell Transplant in Adult Acute Lymphoblastic Leukemia: A Systematic Review and Meta-Analysis": Supplementary Table 1 Search Strategy.docx

| Database | Search Strategy |
| --- | --- |
| Pubmed | ("Whole-Body Irradiation"[Mesh] OR Whole Body Irradiation OR Total Body Irradiations OR Total Body Irradiation OR Whole-Body Irradiations OR Whole-Body Radiation OR Whole Body Radiation OR Whole-Body Radiations OR Radiation, Whole Body) AND ("Precursor Cell Lymphoblastic Leukemia-Lymphoma"[Mesh] OR Precursor Cell Lymphoblastic Leukemia Lymphoma OR Lymphoblastic Leukemia OR Lymphocytic Leukemia, Acute OR Acute Lymphocytic Leukemia OR Leukemia, Acute Lymphocytic OR Leukemia, Lymphocytic, Acute OR Leukemia, Lymphoblastic, Acute OR Lymphoblastic Leukemia, Acute OR Lymphoblastic Lymphoma OR Lymphoma, Lymphoblastic OR Acute Lymphoid Leukemia OR Leukemia, Acute Lymphoid OR Lymphoid Leukemia, Acute OR Leukemia, Lymphoid, Acute OR Leukemia, Lymphoblastic OR Leukemia, Acute Lymphoblastic OR Acute Lymphoblastic Leukemia OR Leukemia, Lymphocytic, Acute, L1 OR ALL, Childhood OR Childhood ALL OR Lymphoblastic Leukemia, Acute, Childhood OR Leukemia, Lymphoblastic, Acute, L1 OR Lymphoblastic Leukemia, Acute, L1 OR Lymphocytic Leukemia, L1 OR L1 Lymphocytic Leukemia OR Leukemia, L1 Lymphocytic OR Leukemia, Lymphocytic, Acute, L2 OR Lymphoblastic Leukemia, Acute, Adult OR Leukemia, Lymphoblastic, Acute, L2 OR Lymphoblastic Leukemia, Acute, L2 OR Lymphocytic Leukemia, L2 OR L2 Lymphocytic Leukemia OR Leukemia, L2 Lymphocytic OR Leukemia, Lymphoblastic, Acute, Philadelphia-Positive) |

Supplementary Table 1: Search Strategy
