## Supplementary material for "Efficacy and Safety of Total Body Irradiation versus Chemotherapy Conditioning for Hematopoietic Stem Cell Transplant in Adult Acute Lymphoblastic Leukemia: A Systematic Review and Meta-Analysis": Supplementary Table 2 Risk of Bias Observational Studies.docx

Risk of bias assessment of cohort studies using Newcastle-Ottawa scale (NOS)

| Study | Selection | | | | Comparability | | Outcome | | | Total |
| --- | --- | --- | --- | --- | --- | --- | --- | --- | --- | --- |
|  | **S1** | **S2** | **S3** | **S4** | **C** | | **O1** | **O2** | **O3** |  |
| Abdeljelil 2024 | * | * | * | * | * | * | * |  | * | 8* |
| Bazarbachi 2020 | * | * | * | * | * | * | * | * | * | 9* |
| Cahu 2015 |  | * | * | * | * |  | * | * | * | 7* |
| Dholaria 2021 | * | * | * | * | * | * | * | * | * | 9* |
| Eroglu 2013 | * | * | * | * | * | * | * | * | * | 9* |
| Greil 2020 | * | * | * | * | * | * | * | * | * | 9* |
| Harada 2019 | * | * | * | * |  |  | * | * | * | 7* |
| Hirschbühl 2023 | * | * | * | * |  |  | * | * | * | 7* |
| Kalaycio 2010 | * | * | * | * |  |  | * |  | * | 6* |
| Kebriaei 2018 | * | * | * | * |  |  | * | * | * | 7* |
| Konuma 2022 | * | * | * | * |  |  | * | * |  | 6* |
| Mora 2024 | * | * | * | * |  |  | * | * | * | 7* |
| Mutsuhashi 2016 | * | * | * | * | * | * | * |  | * | 8* |
| Niu 2022 | * | * | * |  | * | * | * | * | * | 8* |
| Park 2019 | * | * | * |  | * |  | * | * | * | 7* |
| Pavlu 2019 | * | * | * |  | * | * | * | * | * | 8* |
| Sakellari 2018 | * | * | * |  | * | * | * | * | * | 8* |
| Shen 2024 | * | * | * | * | * | * | * | * | * | 9* |
| Swoboda 2022 | * | * | * | * |  | * | * | * | * | 8* |
| Wang 2021 | * | * | * | * |  | * | * | * | * | 8* |
