## Supplementary figures and images for "Efficacy and Safety of Total Body Irradiation versus Chemotherapy Conditioning for Hematopoietic Stem Cell Transplant in Adult Acute Lymphoblastic Leukemia: A Systematic Review and Meta-Analysis"

### Supplementary Figure 1 Risk of Bias in RCTs.png

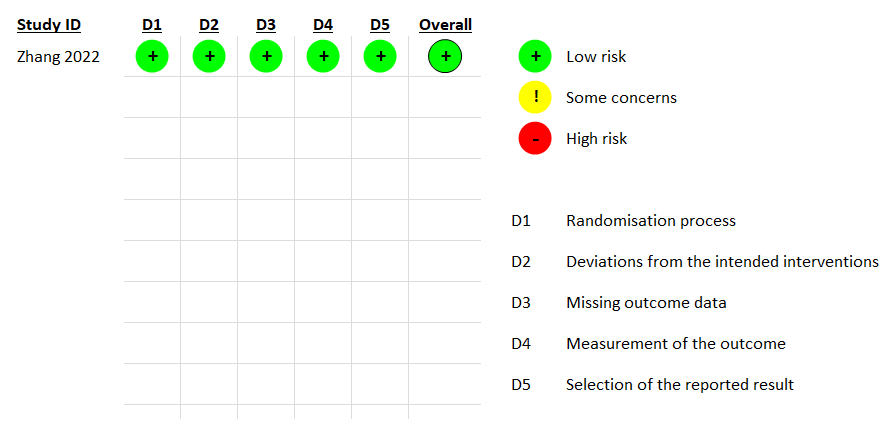

### Supplementary Figure 2 Relapse.png

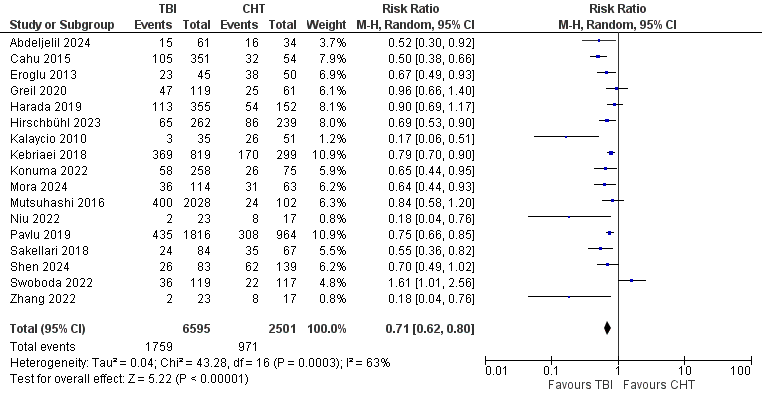

### Supplementary Figure 3 Non-Relapse Mortality.png

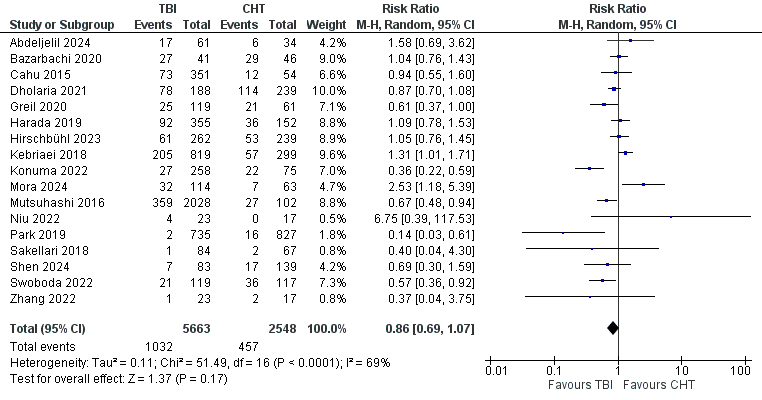

### Supplementary Figure 4 Acute Graft-Versus-Host Disease.png

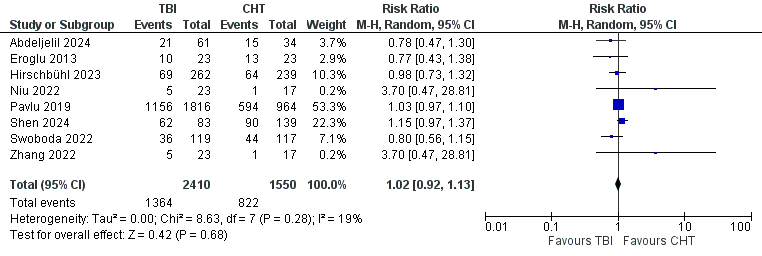

### Supplementary Figure 5 AGVHD Grade 3-4.png

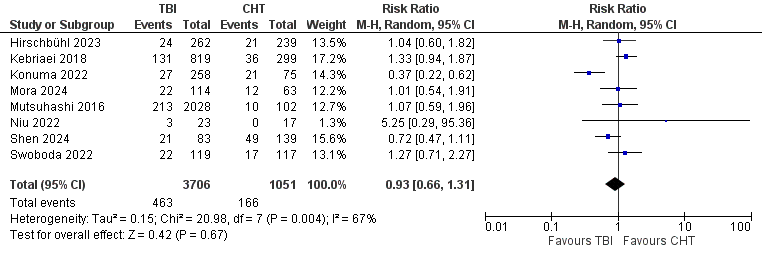

### Supplementary Figure 6 Chronic Graft-Versus-Host Disease.png

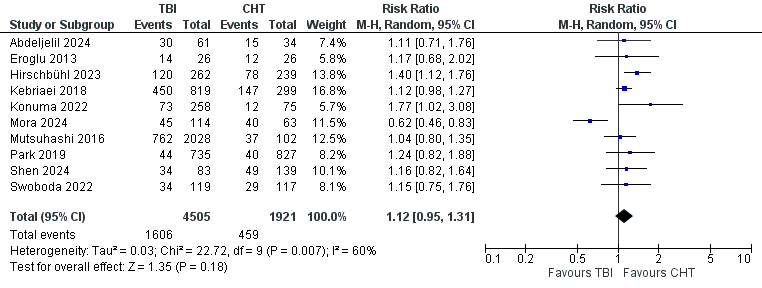
